# Reporting of Pericardial Adverse Events with Immune Checkpoint Inhibitors: A Disproportionality Analysis of the FDA Adverse Event Reporting System

**DOI:** 10.64898/2026.10.19.26365946

**Authors:** Keshav Balakrishna, Vivaan Bhatt, Alessandro Hammond, Michael G. Fradley, Daniel A. Zlotoff, Tomas G. Neilan

## Abstract

**Background:** Immune checkpoint inhibitors (ICIs) can cause immune-related adverse events in almost any organ. While myocarditis has received the most attention, data on pericardial disease with ICI use are limited. We examined pericardial adverse event reporting with nine ICIs using the FDA Adverse Event Reporting System (FAERS).

**Methods:** We combined legacy AERS and current FAERS quarterly files from January 2011 to June 2026 into 18,629,240 unique cases, deduplicated by case number, and removed cases the FDA had listed as deleted. ICI exposure was identified from generic names, brand names, development code names and the active-ingredient field. Pericardial events were defined by seven MedDRA preferred terms. We estimated reporting odds ratios (ROR), proportional reporting ratios (PRR), Bayesian information components (IC) and empirical Bayes geometric means (EBGM). We also examined myocarditis co-reporting, time trends, a comparison with other antineoplastic agents, a MedDRA query-based outcome, and 14 alternative definitions.

**Results:** The analysis included 18,514,045 cases: 277,434 involved an ICI, 34,792 carried a pericardial term, and 1,680 had both. Pericardial events were reported disproportionately with ICIs (ROR 3.35 (95% CI 3.19 to 3.52); PRR 3.34; IC025 1.62). Eight of nine agents had a ROR above 1 (3.32 to 5.63). Among the most frequently reported terms, estimates were highest for cardiac tamponade (5.59 (95% CI 4.98 to 6.28)), followed by pericardial effusion (3.83 (95% CI 3.60 to 4.07)) and pericarditis (2.09 (95% CI 1.89 to 2.31)). After removing cases that also reported myocarditis, the ROR was 3.13 (95% CI 2.98 to 3.30). Most sensitivity analyses gave similar estimates (ROR 3.13 to 3.47); defining exposure by generic drug names alone, which identified only 44% of exposed cases, gave 3.99. Compared with cases naming other L01 antineoplastic agents, the ROR was 1.13 (95% CI 1.08 to 1.19): 1.28 (95% CI 1.20 to 1.36) for ICI-only cases and 0.96 (95% CI 0.89 to 1.04) when another antineoplastic was also named. The ICI-specific reporting odds ratio fell each quarter (odds ratio 0.977 per quarter, 95% CI 0.973 to 0.980).

**Conclusions:** In FAERS, pericardial events were reported more often with ICIs than with other drugs across most agents and terms; however, incidence and causation cannot be determined. The excess over other antineoplastic agents was small, and reporting disproportionality declined over time. These findings support clinical awareness of pericardial disease in patients receiving ICIs and warrant study in data that can measure incidence.

## Background

Immune checkpoint inhibitors (ICIs) are antibodies directed against checkpoint molecules such as cytotoxic T-lymphocyte-associated protein 4 (CTLA-4), programmed cell death protein 1 (PD-1), programmed death ligand 1 (PD-L1) and lymphocyte-activation gene 3 (LAG-3), and are now central to the treatment of many cancers. The same strengthening of the antitumor immune response can also trigger immune-related adverse events (irAEs), which can involve almost any organ [1]. Cardiac irAEs are uncommon but can be severe. Myocarditis has drawn the most attention because of its high reported fatality, whereas pericardial disease, which includes pericarditis, pericardial effusion, cardiac tamponade and, more rarely, constriction, has been less well studied.

Pharmacovigilance and cohort studies have reported a strong association between ICI use and myocarditis [2, 3], and a large analysis of the WHO database also flagged pericardial disease [4]. In a single-center cohort, ICI use was associated with a higher risk of pericarditis or pericardial effusion than was seen in matched patients with metastatic cancer who did not receive ICIs [5]. Two FAERS analyses have examined pericardial events, one directly and one together with myocarditis [6, 7]. However, neither study compared ICIs with other antineoplastic agents, and neither examined how the association of pericardial events with ICI use may have changed over time.

FAERS is a large database of spontaneous reports submitted by manufacturers, health professionals and consumers. Because reporting is voluntary and no denominator of exposed patients is available, FAERS can identify reporting patterns but cannot establish causation or incidence; reporting depends on notoriety, stimulated reporting, drug use, missing information, duplicate reports and confounding by indication and concomitant drugs [8, 9, 10]. We used FAERS, which covers a broad population over many years, to examine reporting patterns of pericardial events with ICIs. We hypothesized that pericardial events would be reported disproportionately more often with ICIs than with other drugs, and we examined whether this excess persisted when ICIs were compared with other antineoplastic agents. We also assessed which agents and terms contributed to the signal, the role of myocarditis co-reporting, and changes over time.

## Methods

### Data source and case definition

The quarterly ASCII files published by the FDA for the legacy Adverse Event Reporting System (AERS; through 2012 Q3) and its successor, FAERS (from 2012 Q4; FAERS replaced AERS in late 2012 and uses a different file structure), were downloaded and processed directly, rather than through the openFDA interface, so that deduplication and the linkage of drug and reaction records remained under local control [8, 9]. The study window opened in 2011 Q1, when ipilimumab reached the market, and closed with 2026 Q2. All 62 archives (seven AERS and 55 FAERS) were present, none was duplicated, and all passed an integrity check. Reporting followed the structure recommended by READUS-PV [11].

The unit of analysis was the case, identified by the FAERS case number (CASEID; CASE in the older files). The two file structures were handled separately and then merged. For AERS, all quarters were pooled and one record was kept per case, selecting the record with the most recent FDA receipt date and breaking ties by report date, manufacturer date, follow-up sequence and ISR. For FAERS, all quarters were pooled and the highest case version was kept for each case number. Drug and reaction records were attached to the retained record, by case number for FAERS and by ISR for AERS. In the AERS files, 146 ISR values were shared by two different case numbers (292 cases), and no drug or reaction records were attached to those cases. Cases present in both systems (70,740) were counted once. The merged dataset held 18,629,240 unique cases: 945,376 from AERS only, 17,613,124 from FAERS only, and 70,740 in both.

Quarterly packages from 2019 Q1 onward include lists of case numbers that the FDA has deleted. The 103,418 cases in the dataset that appeared on these lists were excluded (229,233 distinct case numbers were listed in all). The lists do not cover earlier years, so deletions before 2019 could not be removed.

### Exposure

Each drug name was normalized (upper case, each run of non-alphanumeric characters replaced by one space) and searched for whole-word matches to the generic name, brand name or development code name of nine agents: ipilimumab, tremelimumab, nivolumab, pembrolizumab, cemiplimab, dostarlimab, atezolizumab, durvalumab and avelumab (Appendix A). The active-ingredient field, which is present in the newer FAERS files, was searched in the same way. A drug record counted in any role (primary suspect, secondary suspect, concomitant or interacting), and a record naming several agents counted for each of them. The nine agents are CTLA-4 inhibitors (ipilimumab, tremelimumab), PD-1 inhibitors (nivolumab, pembrolizumab, cemiplimab, dostarlimab) and PD-L1 inhibitors (atezolizumab, durvalumab, avelumab); the nivolumab–relatlimab combination (Opdualag) was captured under nivolumab. Other checkpoint inhibitors (Appendix A), most of which were approved late in the study period, remain investigational, or are marketed mainly outside the United States, were not analyzed as individual exposures. Cases that named one of these other agents and none of the nine study agents (11,777 cases) were excluded, so that they belonged to neither the exposed group nor the reference group. Every remaining case that named none of the nine agents formed the reference group.

### Outcome

Pericardial events were defined by seven MedDRA preferred terms (PTs), matched exactly: pericarditis, pericardial effusion, cardiac tamponade, pericarditis constrictive, autoimmune pericarditis, pericardial disease and immune-mediated pericarditis. To build the list, every PT in the data that contained “pericard” or “tamponade” was reviewed and sorted into one of three groups: a core term; a term added only in sensitivity analyses, such as myopericarditis, pleuropericarditis and the malignant and infective pericardial terms; or a term left out on purpose, namely procedures (for example pericardial drainage) and “bladder tamponade,” which is not cardiac (Appendix B). “Pericardial disease” is a nonspecific term that may overlap with other pericardial diagnoses; it was included in the primary definition and is also reported separately in Table 2. Myocarditis co-reporting was defined by the terms myocarditis, autoimmune myocarditis and immune-mediated myocarditis.

**Table 1.** Disproportionality by agent. The reference group is the 18,236,611 cases naming none of the nine agents. Cases naming several agents appear under each agent. CI, confidence interval; ROR, reporting odds ratio.

|  | Any case naming the agent |  |  | Cases naming only this agent |  |  |
|---|---|---|---|---|---|---|
| Agent | Cases | Events | ROR (95% CI) | Cases | Events | ROR (95% CI) |
| Ipilimumab | 42,189 | 265 | 3.47 (3.08 to 3.92) | 11,518 | 25 | 1.20 (0.81 to 1.77) |
| Tremelimumab | 4,536 | 46 | 5.63 (4.21 to 7.53) | 185 | 0 | not estimable |
| Nivolumab | 95,477 | 711 | 4.12 (3.83 to 4.44) | 64,540 | 443 | 3.80 (3.46 to 4.17) |
| Pembrolizumab | 104,189 | 658 | 3.49 (3.23 to 3.78) | 101,530 | 600 | 3.27 (3.01 to 3.54) |
| Cemiplimab | 3,335 | 20 | 3.32 (2.14 to 5.15) | 3,230 | 11 | 1.88 (1.04 to 3.40) |
| Dostarlimab | 2,256 | 3 | 0.73 (0.24 to 2.27) | 2,234 | 2 | 0.49 (0.12 to 1.97) |
| Atezolizumab | 38,463 | 247 | 3.55 (3.13 to 4.03) | 37,271 | 202 | 3.00 (2.61 to 3.44) |
| Durvalumab | 21,462 | 138 | 3.56 (3.01 to 4.21) | 16,625 | 70 | 2.32 (1.84 to 2.94) |
| Avelumab | 3,976 | 26 | 3.62 (2.46 to 5.32) | 3,794 | 13 | 1.89 (1.10 to 3.26) |

**Table 2.** Disproportionality by preferred term (exposed versus reference cases). IC, information component; IC025 and IC975, lower and upper bounds of its 95% interval. “Pericardial disease” is a nonspecific term that may overlap with other listed diagnoses.

| Preferred term | All cases | Exposed cases | ROR (95% CI) | IC (IC025 to IC975) |
|---|---|---|---|---|
| Pericarditis | 13,059 | 402 | 2.09 (1.89 to 2.31) | 1.03 (0.89 to 1.18) |
| Pericardial effusion | 19,449 | 1,068 | 3.83 (3.60 to 4.07) | 1.87 (1.78 to 1.96) |
| Cardiac tamponade | 3,969 | 311 | 5.59 (4.98 to 6.28) | 2.37 (2.20 to 2.53) |
| Pericarditis constrictive | 338 | 13 | 2.63 (1.51 to 4.58) | 1.20 (0.43 to 1.97) |
| Autoimmune pericarditis | 20 | 12 | 98.60 (40.31 to 241.23) | 3.31 (2.31 to 4.30) |
| Pericardial disease | 222 | 17 | 5.45 (3.32 to 8.94) | 2.05 (1.36 to 2.74) |
| Immune-mediated pericarditis | 52 | 50 | 1643.62 (399.93 to 6755.01) | 4.83 (4.27 to 5.38) |

### Disproportionality analysis

The primary analysis compared, in a 2*×*2 table, the proportion of exposed cases with a pericardial event with the proportion among reference cases (all cases naming none of the nine agents). Because cancer itself can cause pericardial disease, a second comparison restricted the reference group to cases naming other antineoplastic agents (see “Other antineoplastic agents” below). The ROR and PRR were calculated with 95% confidence intervals, together with the Pearson chi-square statistic and the Evans criteria for a signal (PRR of at least 2, chi-square of at least 4, and at least three cases) [10]. The information component (IC) from a Bayesian confidence propagation neural network was also calculated, with a positive lower bound of its 95% interval (IC025) taken as a signal [12], as was the empirical Bayes geometric mean (EBGM) from the gamma-Poisson shrinker, with an EBGM05 (fifth percentile of the posterior distribution) above 2 as the threshold [13]. Because these measures are built from the same counts, they do not provide independent evidence. Agent-level analyses compared cases naming each agent, with or without other agents, with the reference group, and separately cases naming only that agent. Term-level analyses used each preferred term as the outcome in turn.

### Secondary and sensitivity analyses

#### Myocarditis

To determine whether the signal depended on myocarditis, the primary analysis was repeated after removing, from both the ICI-exposed group and the reference group, every case that carried both a pericardial and a myocarditis term. Reports with both kinds of term were also treated as a separate outcome.

#### Time trend

Each case was assigned to the calendar quarter of its earliest FDA receipt date: the initial FDA date for FAERS cases, the date of the retained record for AERS cases, and the earlier of the two for cases in both systems. For each quarter from 2011 Q1 to 2026 Q2, a 2*×*2 table was built. A binomial logistic model for the probability of a pericardial event was then fitted, with terms for quarter, ICI exposure and their interaction, by iteratively reweighted least squares on the grouped counts. The exponentiated interaction coefficient is the change in the ICI-specific reporting odds ratio per quarter. The model was fitted for all quarters and again from 2015 Q1. Cases whose first date fell outside the study window (18,589) were left out of this analysis only.

#### Other antineoplastic agents

To assess whether the excess simply reflects reporting in patients with cancer, ICI cases were compared with cases that named a drug from the WHO Anatomical Therapeutic Chemical (ATC) class L01 (antineoplastic agents) [14] and none of the nine ICIs. The L01 names were taken from the WHO ATC/DDD index (341 distinct names once checkpoint inhibitors were removed), and drug names and active ingredients were searched. ICI cases were split into those naming no L01 agent (“ICI only”) and those naming at least one (“ICI plus L01”), and each group was compared with the L01 group.

#### Standardized MedDRA query

A standardized MedDRA query (SMQ) is a predefined set of MedDRA terms for a medical condition. To allow comparison with earlier work that used an SMQ-based outcome [7], the narrow “Noninfectious myocarditis/pericarditis” SMQ, version 25.0 (14 preferred terms) [15], was applied as an outcome, together with the six pericardial terms within it (autoimmune pericarditis, myopericarditis, pericarditis, pericarditis adhesive, pericarditis constrictive and pleuropericarditis). This differs from the primary outcome in that it uses a published term set rather than the study’s own, and the full query contains many myocarditis terms and is therefore not a pericardial-only outcome.

#### Alternative definitions

To test whether the primary result depended on how cases, exposure or outcomes were defined, fourteen further analyses changed one element of the primary definition at a time (Table 5). They varied the exposure (generic names only, with brand-only cases either kept in or removed from the reference group; suspect roles only; blinded study drugs removed; other checkpoint inhibitors counted as exposed; single-agent cases only), the data source (FAERS era only; AERS era only), the handling of deleted cases (kept), and the outcome (three alternative term sets; malignant and infective terms removed; myocarditis co-reports removed).

### Bayesian shrinkage

The gamma-Poisson shrinker (openEBGM 0.10.0 [16]) was applied to the 63 agent-by-term cells (each of the nine agents paired with each of the seven preferred terms). This empirical Bayes method moves unstable estimates based on few reports toward the null. Expected counts came from the product and event totals in the analysis dataset. The five hyperparameters came from an earlier fit to the full FAERS product-event table and were not re-estimated after the exposure definition was broadened. Posterior percentiles were found by bisection.

### Software and reproducibility

Data processing and the frequentist analyses used Python (pandas and NumPy), and shrinkage used R. Every drug and term list was kept in one configuration file, and every analysis read from a single case-level dataset. Code availability is stated at the end of the paper.

## Results

### Study population

The merged dataset held 18,629,240 unique cases. After removal of 103,418 deleted cases and 11,777 cases that named only a checkpoint inhibitor other than the nine study agents (Appendix A), 18,514,045 cases remained (Figure 1). Of these, 277,434 (1.5%) involved at least one of the nine agents, 34,792 carried a pericardial term, and 1,680 carried both. The remaining 18,236,611 cases named none of the nine agents and formed the reference group; a second comparison restricted to cases naming other antineoplastic agents is presented below. Most exposed cases named a single agent (240,927); 35,222 named two, 957 named three, and 328 named four or more. The most common pairs were ipilimumab with nivolumab (30,039) and tremelimumab with durvalumab (4,342).

**Figure 1.**
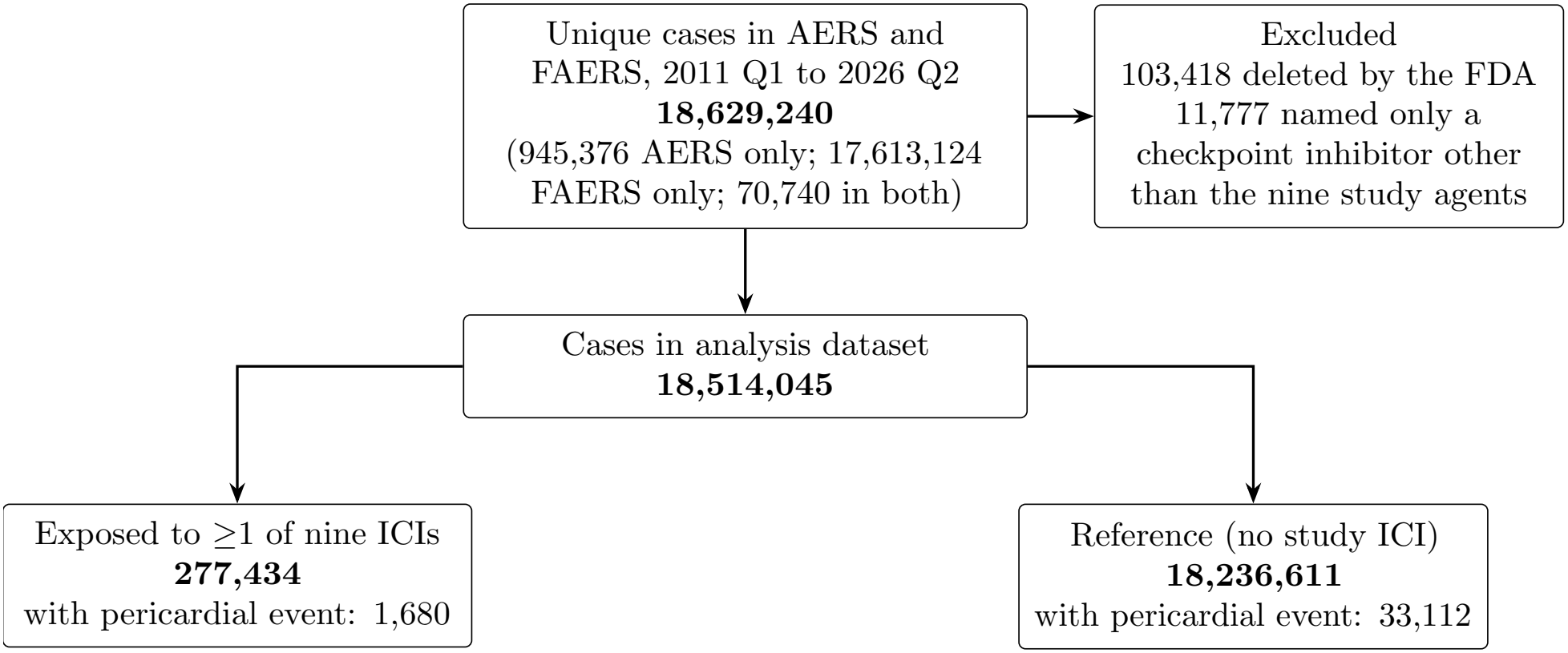
Flow of cases into the analysis.

### Overall disproportionality

Pericardial events were reported for 0.61% of exposed cases (1,680 of 277,434) and 0.18% of reference cases (33,112 of 18,236,611). The ROR was 3.35 (95% CI 3.19 to 3.52) and the PRR 3.34 (95% CI 3.18 to 3.50). The chi-square statistic was 2619.0, and the information component was 1.69 (IC025 1.62).

### Individual agents

Eight of the nine agents had a ROR above 1 (Table 1, Figure 2), with broadly consistent point estimates (3.32 to 5.63). Tremelimumab had the highest point estimate, but 4,342 of its 4,536 exposed cases also named durvalumab, and none of the 185 tremelimumab-only cases had a pericardial event. Ipilimumab showed a similar pattern: its ROR was 3.47 when it was named at all but 1.20 (95% CI 0.81 to 1.77) when it was the only agent (25 events), and 30,039 of its 42,189 exposed cases also named nivolumab. Nivolumab (711 events) gave the most precise estimate among the frequently reported agents. Dostarlimab, with three events, did not meet the Evans criteria.

**Figure 2.**
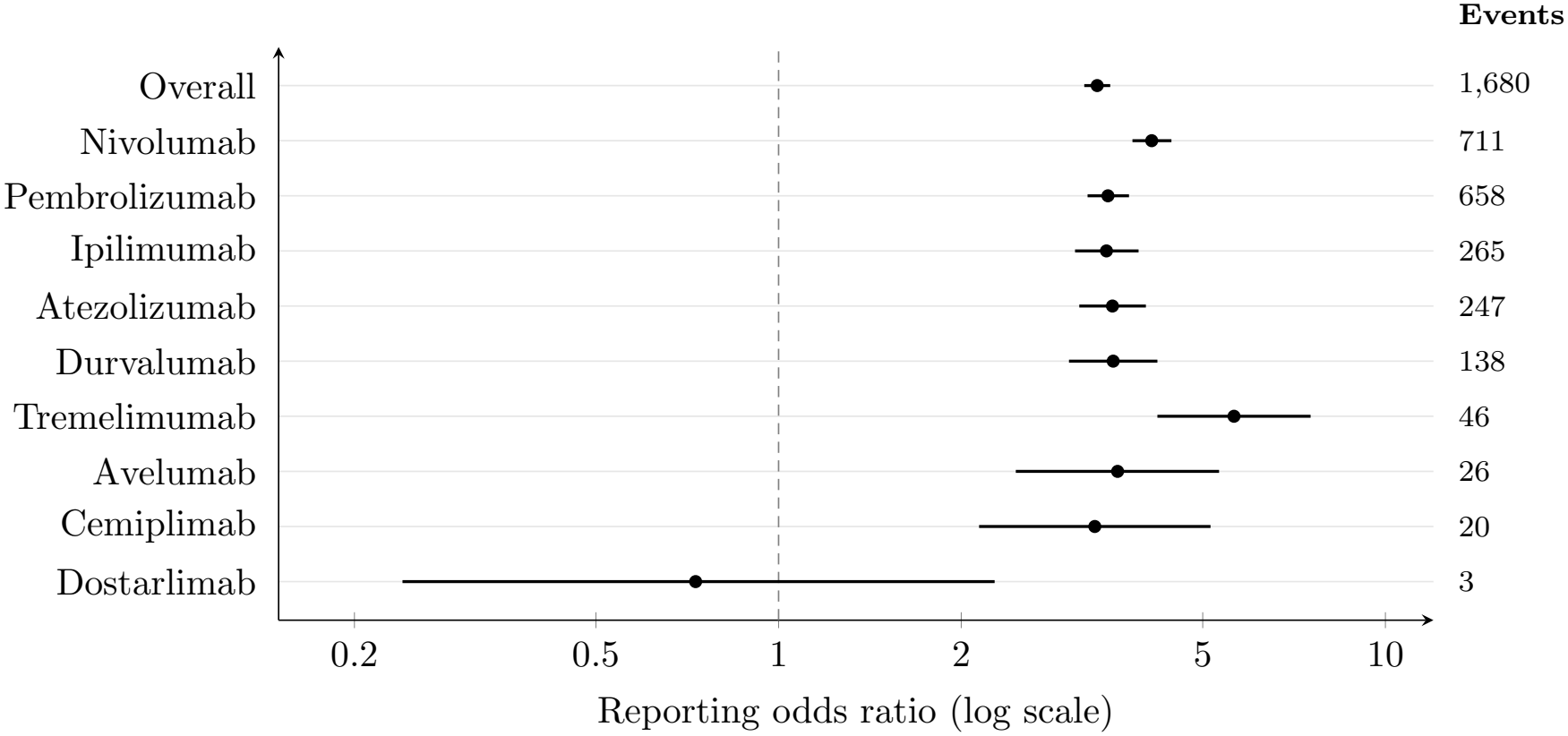
Reporting odds ratios (95% confidence intervals, logarithmic scale) for pericardial events by agent and overall. Numbers to the right are events among cases naming the agent.

### Preferred terms

Every one of the seven terms had a positive lower bound for the information component (Table 2). Pericardial effusion was the most common term among exposed cases, and cardiac tamponade had the strongest ratio among the frequent terms. Autoimmune pericarditis and immune-mediated pericarditis had very large but imprecise ratios because the reference group contained only 8 and 2 cases with these terms. Of all reports of these two terms, 12 of 20 and 50 of 52 involved an ICI.

### Bayesian shrinkage

Of the 63 agent-by-term cells, 46 had at least one exposed event. Twenty-eight cells with at least three events had an EBGM05 above 2 (Table 3 lists the largest). The largest values were for immune-mediated pericarditis with pembrolizumab, nivolumab and ipilimumab. Among the more frequent terms, cardiac tamponade with nivolumab, pembrolizumab and ipilimumab and pericardial effusion with nivolumab, tremelimumab, pembrolizumab, ipilimumab, atezolizumab and durvalumab all had an EBGM05 above 2.9.

**Table 3.** Empirical Bayes estimates for every agent-by-term cell with at least three exposed events and EBGM05 above 2.9 (16 cells). N is the number of exposed cases with the term.

| Agent | Preferred term | N | EBGM | EBGM05 to EBGM95 |
|---|---|---|---|---|
| Pembrolizumab | Immune-mediated pericarditis | 33 | 60.35 | 44.89 to 79.74 |
| Nivolumab | Immune-mediated pericarditis | 14 | 24.77 | 14.58 to 39.51 |
| Ipilimumab | Immune-mediated pericarditis | 9 | 20.40 | 10.22 to 37.85 |
| Nivolumab | Cardiac tamponade | 125 | 5.98 | 5.15 to 6.91 |
| Pembrolizumab | Cardiac tamponade | 130 | 5.71 | 4.93 to 6.58 |
| Nivolumab | Pericardial effusion | 476 | 4.72 | 4.38 to 5.09 |
| Nivolumab | Autoimmune pericarditis | 5 | 9.71 | 4.25 to 21.10 |
| Ipilimumab | Cardiac tamponade | 50 | 5.27 | 4.15 to 6.61 |
| Tremelimumab | Pericardial effusion | 28 | 5.37 | 3.90 to 7.25 |
| Pembrolizumab | Pericardial effusion | 407 | 3.70 | 3.41 to 4.02 |
| Ipilimumab | Pericardial effusion | 163 | 3.64 | 3.20 to 4.13 |
| Nivolumab | Pericardial disease | 9 | 5.66 | 3.18 to 9.48 |
| Ipilimumab | Pericardial disease | 6 | 6.33 | 3.10 to 11.90 |
| Pembrolizumab | Autoimmune pericarditis | 4 | 7.39 | 3.02 to 16.53 |
| Atezolizumab | Pericardial effusion | 140 | 3.43 | 2.98 to 3.93 |
| Durvalumab | Pericardial effusion | 82 | 3.57 | 2.97 to 4.26 |

### Myocarditis co-reporting

Of the 1,680 exposed cases with a pericardial term, 141 (8.4%) also carried a myocarditis term. With those cases removed from both the ICI-exposed group and the reference group, the ROR was

3.13 (95% CI 2.98 to 3.30) (1,539 exposed events). Reports with both kinds of term had a stronger association (ROR 13.78 (95% CI 11.49 to 16.52)).

### Time trend

All 62 quarters from 2011 Q1 to 2026 Q2 contained cases; 54 contained at least one exposed pericardial case, and 41 met the Evans criteria (PRR of at least 2, chi-square of at least 4, and at least three cases). The interaction between ICI exposure and quarter was negative (coefficient *−*0.0238, standard error 0.0019), an odds ratio of 0.977 per quarter (95% CI 0.973 to 0.980), which is about a 2.3% fall in the ICI-specific reporting odds ratio each quarter. Fitted from 2015 Q1 onward, the odds ratio was 0.963 (95% CI 0.959 to 0.967), a fall of about 3.7% per quarter. Pericardial reporting among non-ICI cases rose slightly over the same period (odds ratio 1.008 per quarter).

### Comparison with other antineoplastic agents

We identified 2,616,325 cases that named an antineoplastic agent in ATC class L01 (an “L01 agent”) and none of the nine ICIs, and 13,990 (0.53%) of them carried a pericardial term (Table 4). About 45% of ICI cases also named an L01 agent. Compared with the L01 group, ICI-only cases had a ROR of 1.28 (95% CI 1.20 to 1.36), indicating higher reporting of pericardial events than with other antineoplastic agents, whereas cases naming an ICI and another L01 agent had a ROR of 0.96 (95% CI 0.89 to 1.04). All ICI cases together gave 1.13 (95% CI 1.08 to 1.19).

**Table 4.** Comparison with cases naming another antineoplastic (WHO ATC class L01) agent and no study ICI.

| Group | Cases | Pericardial events | % with event | ROR versus L01 group (95% CI) |
|---|---|---|---|---|
| ICI only | 151,248 | 1,032 | 0.68 | 1.28 (1.20 to 1.36) |
| ICI plus other L01 | 126,186 | 648 | 0.51 | 0.96 (0.89 to 1.04) |
| Any ICI | 277,434 | 1,680 | 0.61 | 1.13 (1.08 to 1.19) |
| Other L01, no ICI (reference) | 2,616,325 | 13,990 | 0.53 | — |

**Table 5.**
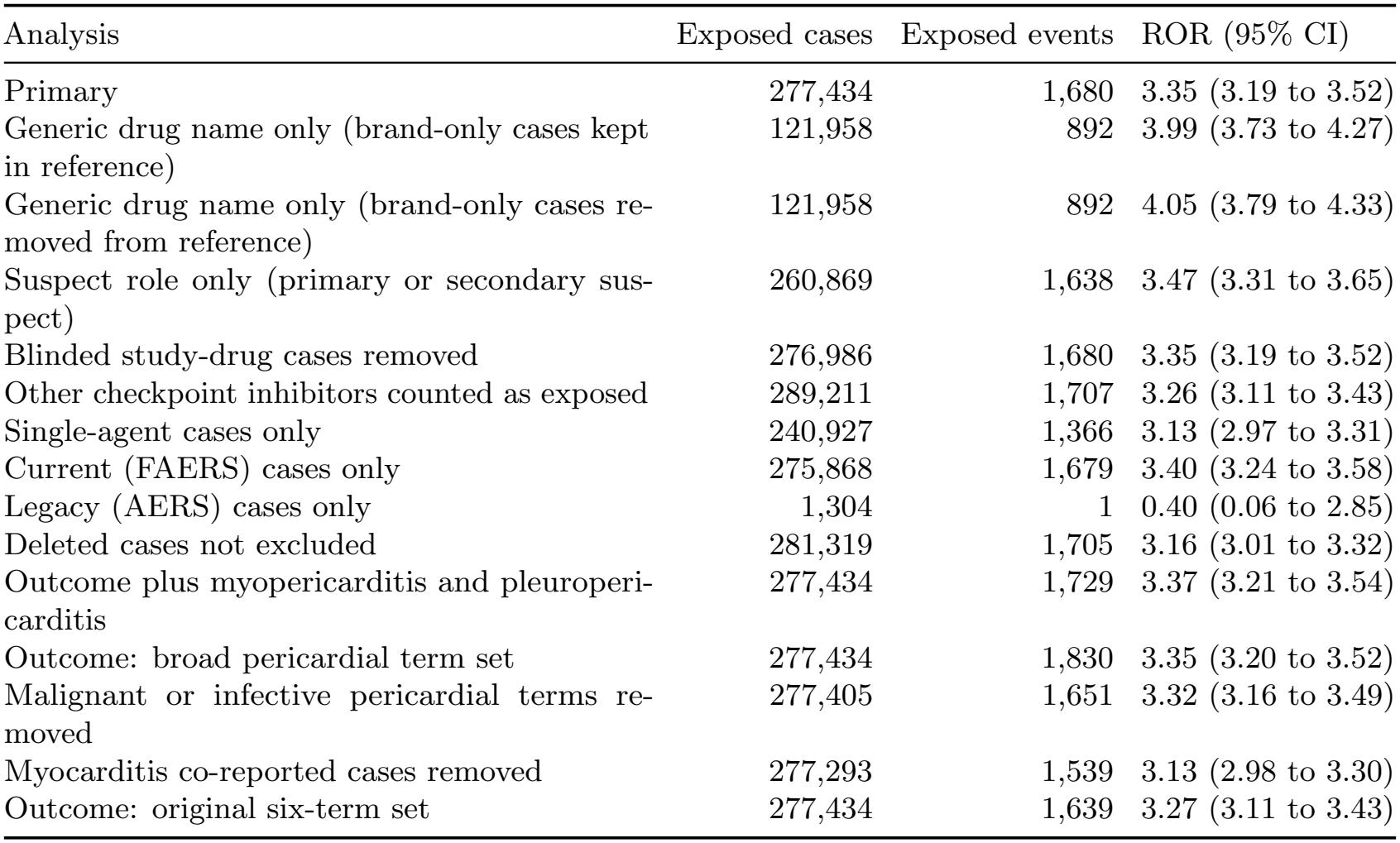
Alternative definitions. Each row changes one element of the primary analysis.

| Analysis | Exposed cases | Exposed events | ROR (95% CI) |
|---|---|---|---|
| Primary | 277,434 | 1,680 | 3.35 (3.19 to 3.52) |
| Generic drug name only (brand-only cases kept in reference) | 121,958 | 892 | 3.99 (3.73 to 4.27) |
| Generic drug name only (brand-only cases re-moved from reference) | 121,958 | 892 | 4.05 (3.79 to 4.33) |
| Suspect role only (primary or secondary sus-pect) | 260,869 | 1,638 | 3.47 (3.31 to 3.65) |
| Blinded study-drug cases removed | 276,986 | 1,680 | 3.35 (3.19 to 3.52) |
| Other checkpoint inhibitors counted as exposed | 289,211 | 1,707 | 3.26 (3.11 to 3.43) |
| Single-agent cases only | 240,927 | 1,366 | 3.13 (2.97 to 3.31) |
| Current (FAERS) cases only | 275,868 | 1,679 | 3.40 (3.24 to 3.58) |
| Legacy (AERS) cases only | 1,304 | 1 | 0.40 (0.06 to 2.85) |
| Deleted cases not excluded | 281,319 | 1,705 | 3.16 (3.01 to 3.32) |
| Outcome plus myopericarditis and pleuroperi-carditis | 277,434 | 1,729 | 3.37 (3.21 to 3.54) |
| Outcome: broad pericardial term set | 277,434 | 1,830 | 3.35 (3.20 to 3.52) |
| Malignant or infective pericardial terms re-moved | 277,405 | 1,651 | 3.32 (3.16 to 3.49) |
| Myocarditis co-reported cases removed | 277,293 | 1,539 | 3.13 (2.98 to 3.30) |
| Outcome: original six-term set | 277,434 | 1,639 | 3.27 (3.11 to 3.43) |

**Table 6.** Study agents and the names searched for each.

| Generic name | Brand and development code names |
|---|---|
| Ipilimumab | bms 734016, bms734016, mdx 010, mdx010, yervoy |
| Tremelimumab | cp 675 206, cp 675206, cp675206, imjudo, ticilimumab |
| Nivolumab | bms 936558, bms936558, mdx 1106, mdx1106, ono 4538, ono4538, opdivo, opdualag |
| Pembrolizumab | keytruda, lambrolizumab, mk 3475, mk3475, sch 900475 |
| Cemiplimab | libtayo, regn 2810, regn2810 |
| Dostarlimab | jemperli, tsr 042, tsr042 |
| Atezolizumab | mpdl3280a, rg 7446, rg7446, tecentriq |
| Durvalumab | imfinzi, medi 4736, medi4736 |
| Avelumab | bavencio, msb 0010718c, msb0010718c |

### Standardized MedDRA query

The full narrow query held 26,654 cases in the analysis dataset and gave a ROR of 15.13 (95% CI 14.66 to 15.61), driven mostly by myocarditis terms (immune-mediated myocarditis alone accounted for 1,380 exposed cases of 1,427). Its six pericardial terms held 14,339 cases and gave a ROR of 2.30 (95% CI 2.10 to 2.52) (484 exposed events).

### Alternative definitions

To test the robustness of the primary result, 14 alternative definitions were each compared with the primary analysis (Table 5). In all analyses other than the two generic-name-only analyses and the legacy (AERS) analysis, the ROR remained between 3.13 and 3.47. The legacy analysis was uninformative because AERS-era cases contained only a single exposed pericardial event. Two changes stood out. First, defining exposure by generic names in the drug-name field alone identified only 44% of exposed cases and gave a higher ROR (3.99 (95% CI 3.73 to 4.27)). Among exposed cases, 0.73% of those identified by generic name had a pericardial event, compared with 0.51% of those it missed. Second, retaining the deleted cases gave a lower ROR (3.16 (95% CI 3.01 to 3.32)). Deleted cases were more likely to carry a pericardial term than retained cases (2.15% against 0.19%), and the difference was concentrated from 2020 onward: from 2021 to 2025 the yearly share among deleted cases ranged from 2.02% to 5.69%, compared with 0.19% to 0.26% among retained cases.

## Discussion

### Summary of findings

In more than 18.5 million cases from FAERS and its predecessor, pericardial events were reported more often with ICIs than with the rest of the database (ROR 3.35). Eight of nine agents had elevated ROR values, with broadly consistent point estimates, and the overall ROR remained largely stable across multiple variations of case, exposure and outcome definitions. Cardiac tamponade (ROR 5.59) and pericardial effusion (ROR 3.83) accounted for most of the cases. Pericarditis was reported less disproportionately (ROR 2.09), and the pericardial terms of the standardized query, which focus on pericarditis, gave a correspondingly lower ROR (2.30). The association persisted after cases that also reported myocarditis were removed (ROR 3.13). In contrast, the excess over other antineoplastic agents was small (ROR 1.28 for ICI-only reports) and absent when another antineoplastic agent was also named (ROR 0.96).

### Comparison with prior work

Our findings extend the earlier FAERS analyses of pericardial events. Ma and colleagues studied reports from January 2011 to March 2020, identified 705 ICI-associated pericardial reports using a list of 15 preferred terms, and found a ROR of 4.87 (95% CI 4.51 to 5.25), with no signal for anti-CTLA-4 agents [6]. They cautioned that the signal might be inflated because pericardial disease is a common complication of cancer. Our ROR of 3.35 should not be set beside theirs as evidence that the signal has weakened. The two studies differ in period, in outcome (their 15-term list included pericardial drainage and calcification), in how exposure was identified, in deduplication, and in the comparison; the time-trend model, fitted within a single dataset, is the better guide to change over time. Yao and colleagues analyzed noninfectious myocarditis and pericarditis together using a standardized MedDRA query in data from 2014 to 2023 (1,112 cases), and reported a ROR of 11.90 (95% CI 11.20 to 12.64) [7]. The estimate for the same query in our data (15.13) was dominated by myocarditis terms and cannot be compared with a pericardial-only estimate. In a single-center cohort with matched patients with metastatic cancer who did not receive ICIs, ICI use was associated with more than a fourfold higher hazard of pericarditis or pericardial effusion (hazard ratio 4.37, 95% CI 2.09 to 9.14) [5]. The excess over other antineoplastic agents in our reporting data was smaller (ROR 1.28 for ICI-only reports), although the two measures are not directly comparable.

### Methodological considerations

Agent-level estimates should be interpreted with caution because reports often named several agents. Tremelimumab had the highest ROR, but almost all of its reports also named durvalumab, and none of the tremelimumab-only reports had a pericardial term. Ipilimumab showed the same pattern: the ROR was 3.47 when it was named at all and 1.20 (95% CI 0.81 to 1.77) when it was the only agent, and most of its reports also named nivolumab. Ma and colleagues likewise found no signal for ipilimumab monotherapy (ROR 1.27, 95% CI 0.84 to 1.93) [6]. Combinations were not analyzed as separate exposures, so whether a combination carries more risk than its components cannot be determined. Nivolumab and pembrolizumab, the most frequently reported agents, gave the most precise estimates (4.12 and 3.49), whereas cemiplimab, avelumab and dostarlimab rested on 20, 26 and 3 events, so their ROR estimates should be interpreted cautiously. The largest ROR values, for autoimmune pericarditis (98.60) and immune-mediated pericarditis (1,644), rested on very small reference groups, and both terms name an immune cause, which is a decision made by the reporter. Most reports of these terms involved an ICI (60% and 96%), so these values reflect how reports are coded once ICI cardiotoxicity is suspected; they are not risk estimates. Finally, the definition of exposure mattered: matching generic names alone identified only 44% of exposed cases and gave a higher ROR (3.99), because reports identified only by a brand name (for example Keytruda) or development code (for example MK-3475) had a lower pericardial share. Studies that rely on generic names alone may therefore overstate a signal of this kind. Excluding FDA-deleted cases, which were enriched for pericardial terms, moved the estimate in the other direction (3.16 with them, 3.35 without them).

### Clinical implications

Most of the signal came from pericardial effusion and cardiac tamponade rather than from pericarditis itself, suggesting that the pericardial events reported with ICIs most often involved fluid accumulation, some of which can be hemodynamically significant. Pericardial effusion is also common in advanced cancer whatever the treatment, and malignant effusion is a particular concern because effusion and tamponade carry most of the signal, as Ma and colleagues also noted [6]. Removing cases with malignant or infective pericardial terms changed the estimate very little (3.32), but those terms identify only a fraction of such cases, and indication data were not used. While the odds of pericardial events were higher in patients who received ICIs than in those receiving other L01 drugs, they were lower than in the comparison with the entire FAERS database. This pattern suggests that part of the signal seen against the full database may reflect the underlying cancer rather than the ICIs themselves. The absence of an excess when another antineoplastic agent was also named (ROR 0.96) may reflect differences between ICI-only and combination reports in cancer type, line of treatment and source of report. These findings support awareness of pericardial disease, including effusion and tamponade, among teams caring for patients receiving ICIs. Additional research is needed to clarify which patients are at greatest risk of ICI-related pericardial events.

The ICI-specific reporting odds ratio fell by about 2.3% per quarter over the whole study period and by about 3.7% per quarter from 2015. Changes in recognition and coding, earlier recognition and management of cardiac toxicity, a growing and more varied ICI-treated population, and a changing mix of reporters could each produce such a decline, and the model cannot tell them apart.

### Limitations

FAERS has no denominator of exposed patients and no information on duration of follow-up, so incidence and absolute risk cannot be estimated. Reporting is voluntary and shaped by notoriety and stimulated reporting, introducing the possibility of reporting bias, and some patient populations may be more likely than others to report adverse events. Some duplicate reports probably remain. Confounding by indication, cancer type, disease stage, other treatment and earlier cardiotoxic exposure could not be adjusted for, and all estimates came from unadjusted 2*×*2 tables. The comparison with other antineoplastic agents only partly addresses confounding by cancer: it depends on name matching against the WHO list, the L01 class includes drugs that are also used outside oncology, such as methotrexate, and the comparison groups probably differ in cancer type, line of treatment and source of report. Exposure depended on drug-name text. Brand and code names were mapped for the nine agents, but the list of other checkpoint inhibitors is not exhaustive, and some names may have been misspelled or missed. Because any drug role counted as exposure, a drug listed only as a concomitant medication counted (a suspect-only analysis gave a similar result). Cases naming several agents contributed to each agent, so the agent-level estimates are not head-to-head comparisons. Legacy AERS cases contributed few exposed events and lack an initial report date, so their quarter reflects the retained record. “Pericardial disease” is a nonspecific term that may overlap with other pericardial diagnoses.

The outcome list and the exposure rules were settled after an initial run of the analysis, which revealed a misspelled preferred term and the shortcomings of generic-name matching, and no protocol was registered in advance. The temporal model assumes a linear trend on the log-odds scale. Shrinkage estimates rest on hyperparameters from an earlier fit that were not re-estimated, and the first mixture component of that fit is degenerate, so EBGM values (particularly for cells with few cases) are secondary and may be too strongly shrunk. Many agent-by-term cells were examined without a formal correction for multiple comparisons. Comparisons with earlier studies are qualitative.

## Conclusions

Pericardial events, particularly pericardial effusion and cardiac tamponade, were reported more often with ICIs than with other drugs in FAERS (ROR 3.35), across most agents and after myocarditis co-reports were removed. The excess over other antineoplastic agents was small (ROR 1.28 for ICI-only reports; 0.96 when another antineoplastic was also named) and reporting disproportionality declined over time. These findings describe reporting patterns, not incidence or causation, but support heightened awareness of pericardial disease among teams caring for patients receiving ICIs and further study to identify the patients at greatest risk.

## Data Availability

Availability of data and materials. FAERS quarterly files are available from the FDA. The analysis code and configuration file are maintained in a public repository (https://github.com/Vivaan-bhatt/faers-pericardial-ici).

https://www.fda.gov/drugs/fda-adverse-event-monitoring-system-aems/fda-adverse-event-monitoring-system-aems-latest-quarterly-data-files

https://fis.fda.gov/extensions/FPD-QDE-FAERS/FPD-QDE-FAERS.html

## Abbreviations

AERS: Adverse Event Reporting System (legacy)
ATC: Anatomical Therapeutic Chemical
CI: confidence interval
EBGM: empirical Bayes geometric mean
FAERS: FDA Adverse Event Reporting System
IC: information component
ICI: immune checkpoint inhibitor
irAE: immune-related adverse event
L01: ATC class for antineoplastic agents
MedDRA: Medical Dictionary for Regulatory Activities
PRR: proportional reporting ratio
PT: preferred term
ROR: reporting odds ratio
SMQ: standardized MedDRA query

## Declarations

### Ethics approval and consent to participate

This study used publicly available, de-identified spontaneous reports from FAERS and its predecessor. It did not involve identifiable human subjects data or any contact with patients, and institutional review board approval was not required.

### Consent for publication

Not applicable.

### Availability of data and materials

FAERS quarterly files are available from the FDA. The analysis code and configuration file are maintained in a public repository (https://github.com/Vivaan-bhatt/faers-pericardial-ici).

## Appendix A. Drug names searched

Names were matched as whole words after normalization (upper case, non-alphanumeric characters replaced by spaces), in drug names and, where present, in active-ingredient fields. Development code names were taken from public drug-approval records and verified at the time of analysis.

### Other checkpoint inhibitors

(cases naming only these were excluded from the primary analysis): retifanlimab, zynyz, toripalimab, loqtorzi, tislelizumab, tevimbra, cosibelimab, unloxcyt, penpulimab, relatlimab, sintilimab, tyvyt, camrelizumab, sugemalimab, serplulimab, envafolimab, zimberelimab, cadonilimab, spartalizumab, balstilimab, prolgolimab.

### Blinded study drugs

Records containing the words “blinded”, “blind” were flagged for a sensitivity analysis.

## Appendix B. MedDRA preferred terms

### Primary outcome (7 terms)

pericarditis, pericardial effusion, cardiac tamponade, pericarditis constrictive, autoimmune pericarditis, pericardial disease, immune-mediated pericarditis.

### Added in the immune-focused sensitivity analysis

myopericarditis, pleuropericarditis.

### Added in the broad sensitivity analysis

(all non-procedural pericardial terms in the data; the terms marked with an asterisk are the malignant, infective and other-cause terms removed in one analysis): pericardial haemorrhage, pneumopericardium, pericardial fibrosis, pericarditis adhesive, pericardial calcification, intrapericardial thrombosis, pericardial mass, pericardial cyst, pericardial rub, postpericardiotomy syndrome, pericardial rupture, pericardial injury, pericardial perforation, pericardiocutaneous fistula, pericardial lipoma, benign pericardium neoplasm, camptodactyly-arthropathy-coxa vara-pericarditis syndrome, pericardial effusion malignant*, pericarditis malignant*, pericardial neoplasm*, malignant pericardial neoplasm*, pericardial mesothelioma malignant*, pericardial mesothelioma malignant recurrent*, pericarditis infective*, infective pericardial effusion*, viral pericarditis*, purulent pericarditis*, bacterial pericarditis*, pericarditis tuberculous*, pericarditis fungal*, coxsackie pericarditis*, pericarditis mycoplasmal*, cytomegalovirus pericarditis*, pericarditis meningococcal*, pericarditis histoplasma*, atypical mycobacterium pericarditis*, radiation pericarditis*, pericarditis uraemic*, pericarditis rheumatic*, pericarditis lupus*.

### Excluded on purpose

pericardial drainage, pericardial excision, pericardiotomy, pericardial operation, pericardial cyst resection, biopsy pericardium abnormal, pericardial decompression syndrome, bladder tamponade.

### Myocarditis terms

myocarditis, autoimmune myocarditis, immune-mediated myocarditis.

### Narrow Noninfectious myocarditis/pericarditis query, version 25.0 (14 terms)

autoimmune myocarditis, autoimmune pericarditis, carditis, chronic myocarditis, eosinophilic myocarditis, giant cell myocarditis, hypersensitivity myocarditis, immune-mediated myocarditis, myocarditis, myopericarditis, pericarditis, pericarditis adhesive, pericarditis constrictive, pleuropericarditis. The six pericardial terms within it are autoimmune pericarditis, myopericarditis, pericarditis, pericarditis adhesive, pericarditis constrictive, pleuropericarditis.

